# Noninvasive estimation of mean pulmonary artery pressure by CMR in under 2 minutes scan time

**DOI:** 10.1101/2023.04.04.23288073

**Authors:** Goran Abdula, Joao G Ramos, David Marlevi, Alexander Fyrdahl, Henrik Engblom, Peder Sörensson, Daniel Giese, Ning Jin, Andreas Sigfridsson, Martin Ugander

**Affiliations:** Department of Clinical Physiology, Karolinska University Hospital, and Karolinska Institutet, Stockholm, Sweden; Institute for Medical Engineering and Science, Massachusetts Institute of Technology, Cambridge, Massachusetts, USA; Department of Cardiology, Karolinska University Hospital, and Karolinska Institutet, Stockholm, Sweden; Siemens Healthcare, Germany; Siemens Medical Solutions, Cleveland, Ohio, USA; Kolling Institute, Royal North Shore Hospital, and University of Sydney, Sydney, Australia

**Keywords:** pulmonary hypertension, magnetic resonance imaging (MRI), four-dimensional flow, accelerated imaging

## Abstract

**Background:** Non-invasive estimation of mean pulmonary artery pressure (mPAP) by cardiovascular magnetic resonance (CMR) four-dimensional (4D) flow analysis has shown excellent agreement with invasive right heart catheterization. However, clinical application is limited by relatively long scan times.

**Objectives:** The aim of this study was to evaluate the accuracy and time reduction of compressed sensing (CS) accelerated acquisition for mPAP estimation.

**Methods:** Patients (n=51) referred for clinical CMR at 1.5T or 3T underwent imaging with both a prototype CS□accelerated and a non-CS-accelerated flow sequence acquiring time-resolved multiple 2D slice phase contrast three-directional velocity-encoded images covering the pulmonary artery. Prototype software was used for blinded analysis of pulmonary artery (PA) vortex duration to estimate mPAP as previously validated.

**Results:** CS-accelerated and non-CS-accelerated acquisition showed increased mPAP in 22/51 (43%) and 24/51 (47%) patients, respectively. Mean bias for estimating mPAP between the two methods was 0.1±1.9 mmHg and the intraclass correlation coefficient was 0.97 [95% confidence interval 0.94-0.98]. Effective scan time was lower for the CS-accelerated acquisition (1 min 55 sec ± 27 sec vs 9 min 6 sec ± 2 min 20 sec, p<0.001, 79% reduction).

**Conclusions:** CS-accelerated CMR acquisition enables preserved accuracy for estimating mPAP compared to a non-CS-accelerated sequence, allowing for an average scan time of less than 2 minutes. CS-acceleration thereby increases the clinical utility of CMR 4D flow analysis to estimate mPAP.

## Introduction

Pulmonary hypertension (PH) is a progressive disease associated with high mortality regardless of the underlying pathophysiological etiology (1). PH also has a dominant effect on healthcare worldwide, with about 1% of the global population, up to 10% of individuals aged over 65 years, and at least 50% of patients with heart failure (HF) all suffering from PH (2). To date, diagnostic evaluation of PH is determined by invasive right heart catheterization (RHC), defined by a mean pulmonary artery pressure (mPAP) of 20 mmHg or greater at rest (3). However, the invasive nature of RHC procedures limits applicability, as well as use for early diagnostic screening of PH. Instead, non-invasive imaging, and cardiovascular magnetic resonance (CMR) in particular, offers an important role in the clinical management of patients with PH (4), providing accurate and reproducible assessment of left ventricular (LV) and right ventricular (RV) volumes, function, and mass. CMR 4D flow analysis has also been proposed as a potential non-invasive tool for estimating mPAP (5, 6), empirically relating the persistence of a pathological flow vortex in the main pulmonary artery to mPAP. Within this space, 4D flow analysis has shown excellent agreement with invasive right heart catheterization and superior diagnostic accuracy as compared to Doppler echocardiography for estimating mPAP (7). However, clinical application of the method is limited in part by relatively long scan times, with previously utilized protocols requiring approximately 10 minutes of acquisition for complete flow mapping of the main pulmonary artery. Current developments in acceleration techniques such as compressed sensing (CS) are promising and can potentially reduce the scan times. However, the accuracy of 4D flow analysis for CS-accelerated compared to non-CS-accelerated acquisition is not known. Therefore, the aim of this study was to perform a head-to-head comparison of CS-accelerated and non-CS-accelerated flow acquisition, assessing discrepancies in estimated mPAP as well as effective scan time.

## Methods

### Patient population

To evaluate the differences between flow sequences with regards to non-invasive mPAP estimation, a prospective study protocol was defined. Between May 2020 and January 2021, n = 55 consecutive patients referred for clinical CMR were enrolled in this study. The cohort size was based on power calculations seeking to infer a possible difference of 6% in PA vortex duration between sequences, using on a previously known mean vortex duration of approximately 10 ± 10% in an average clinical population. Sample size calculation for α=0.05, β=0.20, and power of 0.80 was n=44. Inclusion criteria included patients referred for clinical CMR on diagnostic basis. Exclusion criteria included known contraindications for CMR as well as arrhythmia, valve prosthesis, or known pulmonary valvular disease. Patient demographics and clinical data of the study population are summarized in Table 1. The study was approved by the Stockholm Regional Board of ethics committee, and written informed consent was obtained from each patient before enrollment in the study (EPN: 2011/1077-31/3).

**Table 1.** Summary of Patients Demographics and Clinical Characteristics

| Patients | PH n=24 | Non-PH n=27 | p-value |
| --- | --- | --- | --- |
| Age, years | 49±17 | 49±19 | 0.974 |
| Female sex, % | 33% | 22% | 0.666 |
| BSA, m<sup>2</sup> | 1.95±0.25 | 1.93±0.20 | 0.799 |
| HR, bpm | 67±14 | 68±10 | 0.975 |
| Clinical Diagnosis |  |  |  |
| IHD, n (%) | 2 (8) | 5 (19) | 0.659 |
| CMP, n (%) | 6 (25) | 6 (22) | 0.964 |
| VHD, n (%) | 3 (13) | 1 (0.4) | 0.770 |
| PH, n (%) | 3 (13) | 0 (0) | 0.709 |
| Myocarditis, n (%) | 4 (17) | 4 (15) | 0.969 |
| Normal CMR, n (%) | 6 (2) | 11 (41) | 0.844 |

BSA; Body Surface Area, bpm; Beats Per minute, IHD; Ischemic Heart Disease, VHD; Valvular Heart Disease, CMP; Cardiomyopathy, PH; Pulmonary Hypertension, CMR; Cardiac Magnetic Resonance

### CMR protocol

CMR was performed at either 1.5 T (n = 21) or 3 T (n = 34) (MAGNETOM Aera or Skyra, Siemens Healthcare, Erlangen, Germany) using phased array receiver coils with electrocardiography (ECG) gating. As part of the clinical routine, the examination included standard breath-hold cine imaging with steady-state free precession (SSFP) in the short axis (SA) and the standard long axis planes. Hemodynamic flow mapping was acquired by positioning a stack of 6-10 gapless slices covering the main pulmonary artery and the right ventricular outflow tract using multiple 2D slices of ECG-gated time-resolved phase contrast with three-directional velocity encoding, with either GRAPPA-acceleration (R=2) with 22 integrated calibration lines, denoted M2D, or CS acceleration with dual-density phase-encoding using a prototype sequence, denoted CS-M2D. The undersampling factor was R=3 in the central region and R=11 in the peripheral region, resulting in an effective acceleration factor of R=7.7. The relevant parameters for the CS reconstruction were as follows; 40 iterations, spatial regularization factor λ_S_ = 0.0003 and temporal regularization factor *λ*_t_ = 0.001.

The two phase-contrast sequences were acquired in direct succession using identical geometry and in free breathing. Common parameters for both sequences were velocity encoding direction 90 cm/s in all three spatial directions, three-fold averaging to suppress breathing artifacts, in-plane field of view 340 × 276 mm^2^, k-space matrix size 192 × 138 corresponding to an acquired in-plane pixel size of 1.8 × 2.0 mm^2^, slice thickness 6 mm, bandwidth 450 Hz/px, flip angle 15°. Both sequences used retrospective cardiac gating, reconstructing to 20 interpolated time frames.

M2D was acquired using TE/TR 4.1/6.4 ms, resulting in an acquired temporal resolution of 77 ms. CS-M2D was acquired using TE/TR 4.1/6.3 ms, resulting in an acquired temporal resolution of 75 ms.

### CMR image processing and image-based mPAP estimation

All flow datasets were de-identified and analyzed using prototype software (4D Flow, Siemens Healthcare, Erlangen, Germany). A fully automatic pre-processing step was applied that performed eddy current compensation and phase-unwrapping as needed. The right ventricular outflow tract and pulmonary artery were manually segmented to capture the primary flow features in these areas. Streamlines were utilized to qualitatively visualize the presence of potential PA vortices. For quantitative assessment, vector visualization was subsequently used while seeding in 2D views at full reconstructed voxel resolution in order to detect flow vortices in the main pulmonary artery, following methodologies described in previous work (8). Briefly, the presence of a vortex was determined by identifying closed concentric rings in the 4D velocity vector field within the main pulmonary artery. Vortex presence was expressed in terms of number of time frames where a vortex could be detected and converted into percentages out of the total number of frames in the cardiac cycle. The complete process is illustrated in Figure 1. Mean pulmonary arterial pressure (mPAP) was then estimated using a previously described (9) empirically determined equation stating:

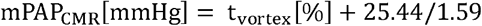

**Figure 1.**
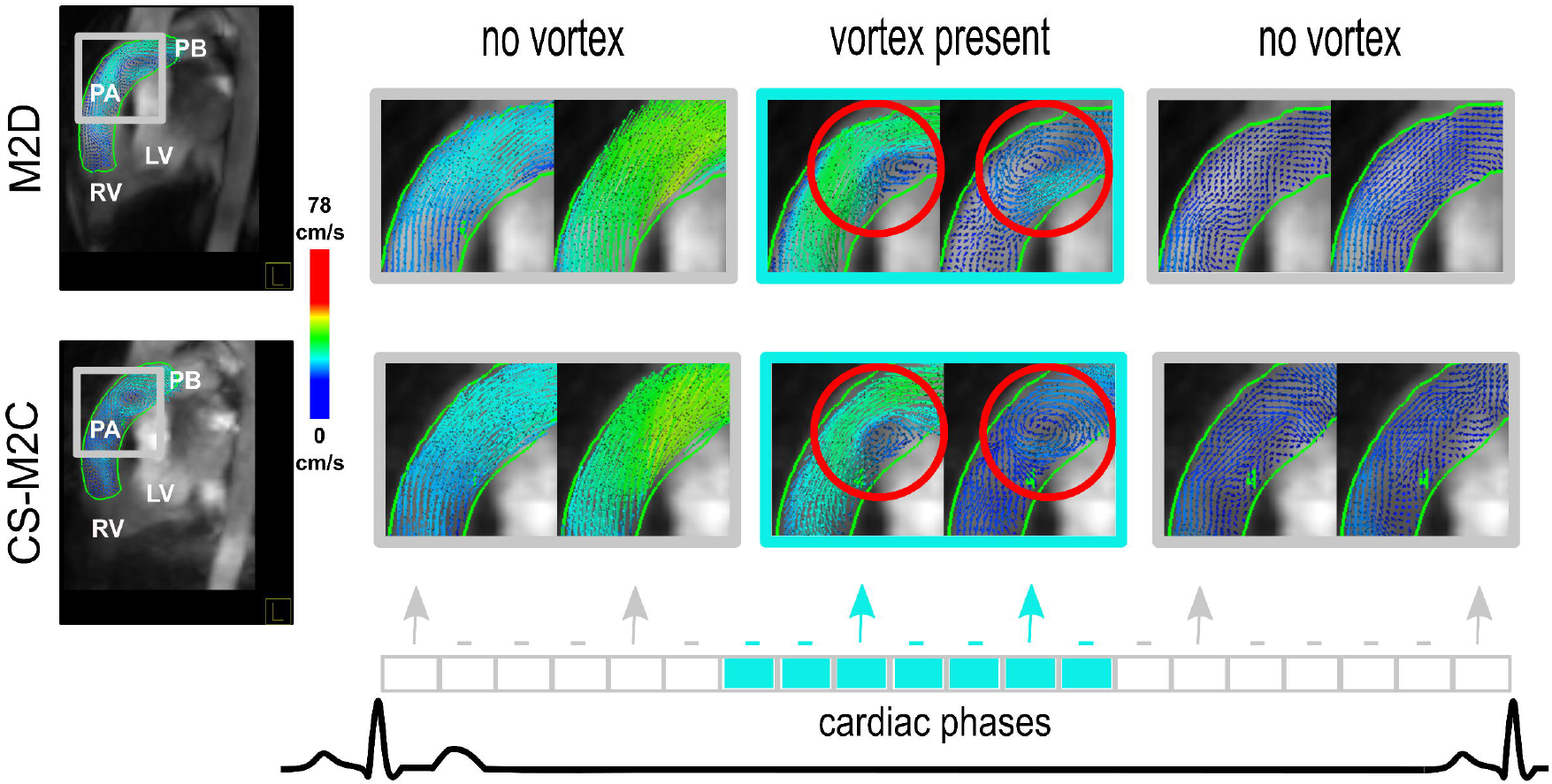
Matched images from selected time frames from a representative patient showing a velocity vector field multiplanar reformat using M2D (top panel) and CS-M2D (bottom panel) multiple 2D slice three-directional time-resolved phase contrast velocity encoded sequences, respectively. Boxes in grey represent successive time frames in the cardiac cycle. The boxes highlighted in light blue represent the time frames in which a vortex could be visualized. M2D – Multiplanar 2D, CS-M2D – Compressed Sensing Multiplanar 2D, RV – right ventricle, LV – left ventricle, PA – main pulmonary artery, PB – pulmonary bifurcation.

The 4D flow analysis was performed by two independent readers (GA and JGR). The analysis was performed interchangeably on both M2D and CS-M2D datasets in a in randomized fashion.

### Statistical analysis

Statistical testing was performed using freely available analysis software (RStudio 1.4, Boston, MA, USA). Continuous variables were expressed as mean ± standard deviation (SD) and categorical variables were presented as percentages. Differences in mean mPAP between CS-M2D and M2D estimations were analyzed by student’s t-test. The Wilcoxon signed-rank test was used to compare the scan time duration for M2D and CS-M2D. The relationships between mPAP derived from M2D vs. CS-M2D were investigated using the Spearman rank correlation coefficient and Bland-Altman analysis. Likewise, regression analysis was performed to quantify differences in mPAP estimation using CS-M2D and M2D across measurements. Possible bias between field strengths (1.5T vs. 3T) was analyzed using an unpaired t-test. Furthermore, interobserver variability was evaluated by performing Bland-Altman analysis and calculating the intraclass correlation coefficient (ICC) and 95% confidence interval (95%CI) of a randomly selected set of n=20 patients. A p-value<0.05 was considered statistically significant.

## Results

### Demographics and baseline characteristics

A total of 55 patients were consecutively recruited and 4 patients were excluded due to poor image quality (two excluded based on the CS-M2D, one based on the M2D, and one due to unrecoverable phase aliasing). As such, 51 patients were included with both CS-M2D and M2D images successfully acquired, with data eligible for 4D flow analysis and estimation of mPAP.

#### Diagnostic differentiation of pulmonary hypertension

Out of the total of 51 patients, analysis by M2D indicated increased mPAP (>20 mmHg) in 24 (47%) patients, and normal evaluated mPAP (≤20 mmHg) in 27 (53%) patients. By comparison, CS-M2D identified 22 (43%) patients with increased, and 29 (57%) patients with normal mPAP, respectively. As such, discrepancies between M2D and CS-M2D in diagnostic differentiation was observed in two patients. Note that in these patients, both estimates were very close to the clinical threshold (mPAP=22 mmHg by M2D vs. mPAP=19 mmHg by CS-M2D). Furthermore, in the patient group with mPAP<20 mmHg by M2D (n=27), 22 of these exhibited no visible vortex in any of the flow sequences, 4 patients had a pathological vortex formation in the main pulmonary artery identified only by CS-M2D, and one patient had pathological vortex formation in the main pulmonary artery identified only by M2D.

#### Quantitative mPAP estimation

Overall, estimation of mPAP between M2D and CS-M2D did not differ (21.7±7.0 vs 21.9±7.2 mmHg, p=0.641). In particular, as shown in Figure 2A, mPAP estimated by CS-M2D revealed excellent agreement with estimates obtained by M2D (R^2^=0.93, P=0.001), and with negligible bias (mean bias 0.2±2.3 mmHg at 1.5 T (n=20), 0.1±1.5 mmHg at 3 T; overall mean bias 0.1±1.9 mmHg (Figure 2B).

**Figure 2.**
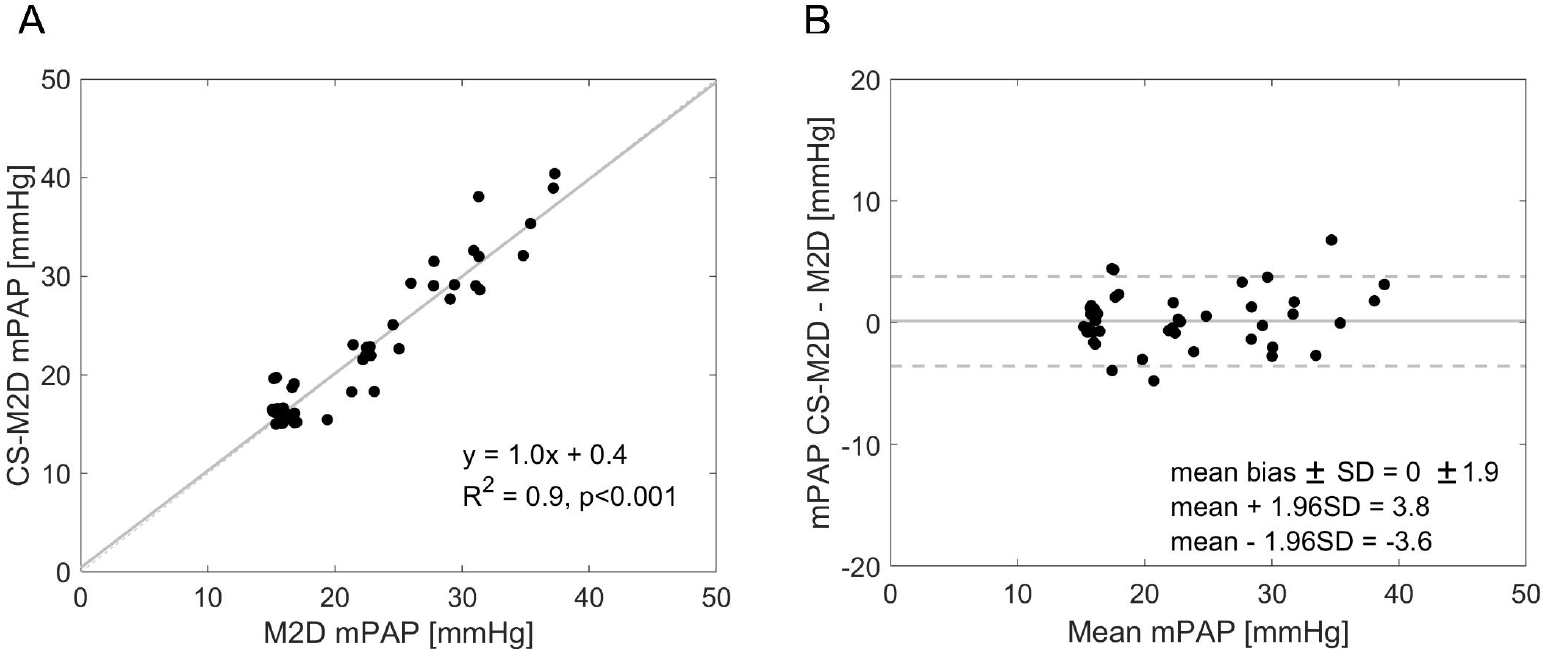
(A) Linear regression (solid line) of estimated mean pulmonary artery pressure (mPAP) by compressed sensing (CS)-accelerated and conventional 4D flow analysis in all analysed patients. Dotted line shows line of identity. (B) Bland-Altman plot of estimated mPAP by 2D conventional and CS CMR flow in patients with both observable vortex by 4D flow analysis. Mean±SD bias was 0.1±1.9 mmHg. Note that data jitter is introduced to improve visibility of overlapping data points. M2D – Multiplanar 2D, CS-M2D – Compressed Sensing Multiplanar 2D

#### Effective scan time

Across all subjects, scan time was 9 min 6 sec ± 2 min 20 sec for M2D compared to 1 min 55 sec ± 27 sec for CS-M2D, representing a reduction of 79% in effective scan time (p<0.001).

### Interobserver variability

From the interobserver variability analysis, good agreement was observed with the intraclass correlation coefficient (ICC) for the estimation of mPAP being 0.97 [95%CI 0.94-0.98]. Similarly, the overall interobserver variability yielded a good agreement for mPAP by both the M2D and CS-M2D, with ICC of 0.95 [95%CI 0.89-0.98] for estimated mPAP. As shown in Figure 3, high linear regression coefficients and low mean bias were also all reported for interobserver variability analysis on both CS-M2D and M2D analysis, respectively.

**Figure 3.**
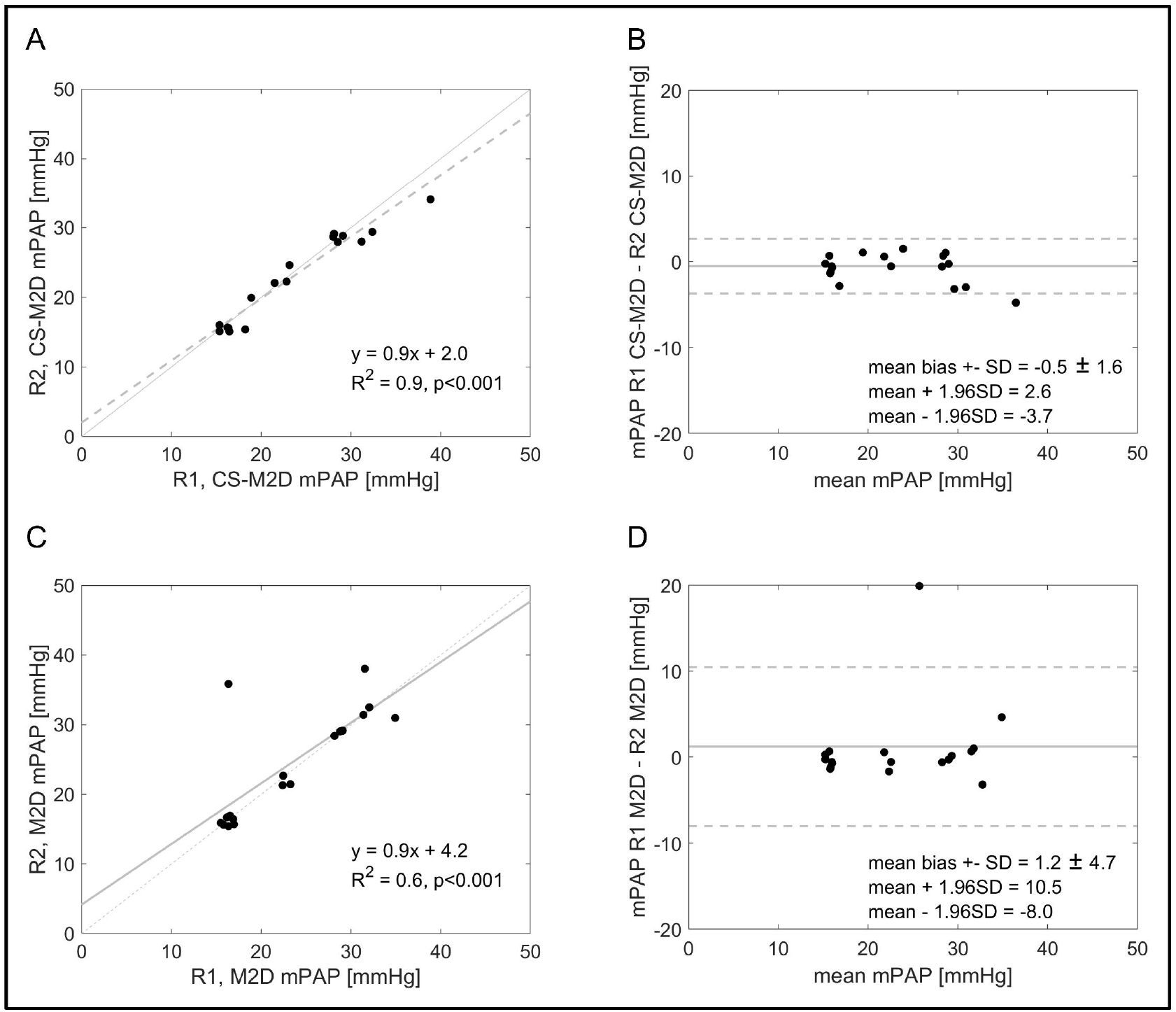
Linear regression and Bland-Altman plots of mean pulmonary artery pressure (mPAP) estimated by Reader 1 (R1) and Reader 2 (R2) by Compressed Sensing Multiplanar 2D (CS-M2D) (A,B) and Multiplanar 2D (M2D) (C,D). Note that data jitter is introduced to improve visibility of overlapping data points.

## Discussion

The main finding of the study is that CS-accelerated CMR acquisition enables preserved accuracy for estimating mPAP compared to a non-CS-accelerated sequence, allowing for an average scan time of less than 2 minutes. This represents a substantial improvement opening up for non-invasive mPAP estimation without extensively added acquisition duration for existing clinical protocols. The clinical potential of CS-M2D is further emphasized by the minimal bias observed between different field strengths (1.5 T vs. 3.0 T) and different clinical readers.

In the current study, estimation of mPAP from phase-contrast flow images was based on the empirical regression model established by Reiter et al (9), with image-derived vortex duration shown to carry high diagnostic accuracy for the estimation of pathological mPAP exceeding 16 mmHg. These results were also corroborated by a recent study in which M2D exhibited excellent agreement with invasively measured mPAP (n=40) and superior diagnostic performance for detecting PH compared to Doppler echocardiography (7). The results of the current study represent a continuation towards routine clinical implementation, incorporating sequence acceleration through CS protocols, with resulting scan time efficiency and maintained accuracy that pushes the utility towards a more streamlined clinical workflow.

Within the setting of PH, RHC is regarded as the reference standard for assessing mPAP and establishing the diagnosis. However, the invasiveness, complication risk (11), and relatively long procedure time associated with percutaneous catheterization limit its use for screening purposes and may delay diagnosis (12). Instead, Doppler echocardiography remains the primary non-invasive alternative technique used to assess patients with suspected PH, where simplified assessment of the apparent hemodynamic environment is used to estimate peak systolic pulmonary pressure (13). However, echocardiographic assessment of pulmonary pressure has several limitations including limitations in acoustic window (14) and accuracy (15), as well as a modest correlation between systolic PA and mPAP (16). Moreover, almost half of all patients being referred for RHC lack a detectable tricuspid regurgitation jet (TR) by Doppler imaging (17), effectively reducing the applicability of echocardiography. As a result, there is still a clear clinical need for alternative and precise non-invasive methods for the clinical estimation of mPAP, moving beyond surrogate estimates from right ventricular (RV) structure and function. As shown in the current study, CS-accelerated phase-contrast CMR could represent such a tool, effectively enabling accurate mPAP estimation without any substantial addition in protocol time. CMR adds incremental diagnostic value for patients with suspected PH, with standardized sequences allowing for simultaneous quantification of LV and RV function, myocardial fibrosis or other underlying conditions, all in the very same imaging session (18). In the clinical setting, CMR shortens the time delay for PH diagnosis and treatment which has the potential to improve prognosis in PH.

### Limitation

There are some limitations that should be acknowledged. A relatively small study population was utilized. Although sufficiently powered to identify the expected differences in mPAP between the compared sequences, this does not mean that the current data necessarily can be generalized to a wider spectrum of cardiac conditions. Specifically, we chose to exclude patients with arrhythmia and severe pulmonary valvular disease, since these conditions are known to interfere with phase-contrast CMR data quality. The ability to non-invasively estimate mPAP by CMR in these patients – and in particular the difference between M2D and CS-M2D – would thus have to be assessed in separate, future analysis. Our study utilized intramodality validation, with the non-CS-accelerated M2D sequence used as reference for the CS-M2D equivalent. This was performed in light of previous work successfully validating M2D analysis against invasive catheterization at multiple institutions (5, 6). Nevertheless, head-to-head comparison between CS-accelerated CMR and invasive RHC could be used to corroborate the findings of the current study. Although we reported a sizeable reduction in acquisition time for CS-M2D vs M2D, this did not include the time required for post-processing and quantitative estimation of mPAP between the two methods. However, although minor qualitative differences were indicated between sequences by the clinical readers, the time required to manually assess vortex presence was identical between the two sequences, especially when considering the same patient and scan session (data not shown).

## Conclusions

CS-accelerated CMR flow acquisition for 4D flow analysis exhibits excellent agreement with non-CS-accelerated acquisition for quantitative estimation of mPAP. Further, the CS-accelerated sequence reduced effective scan time to under 2 minutes, and was robust at both acquisition field strengths and inter-reader variations. Consequently, CS-accelerated CMR in the diagnosis of PH shows clinical utility.

## Data Availability

All data produced in the present work are contained in the manuscript

## Abbrevations and Acronyms

PH: Pulmonary hypertension
mPAP: Mean pulmonary artery pressure
HF: Heart failure
RHC: Right heart catheterization
LV: Left ventricle
RV: Right ventricle
CS: Compressed sensing

## Clinical perspectives

### Clinical Competencies

The CS-accelerated M2D flow extends the use of CMR 4D flow analysis to estimate mPAP noninvasively.

### Translational Outlook

The accuracy of the CS-accelerated M2D flow technique among different types of pulmonary hypertension needs to be verified in a prospective multicenter study. Additionally, to evaluate the impact of non-invasive assessment of mPAP on diagnosis and clinical management of patients with pulmonary hypertension.

## Acknowledgments

GA participated in the study design, performed data analysis, performed statistical analysis, and drafted the manuscript. JGR participated in study design and data analysis and interpretation. DM participated in study design and data analysis and interpretation. AF, AS, DG and NJ participated in study design, and participated in data acquisition and interpretation. HE and PS participated in study design, data interpretation, and manuscript revision. MU designed the study, participated in data analysis and interpretation, and helped to draft the manuscript. The final manuscript was read and approved by all authors.

## Figure titles and legends

**Central illustration.**
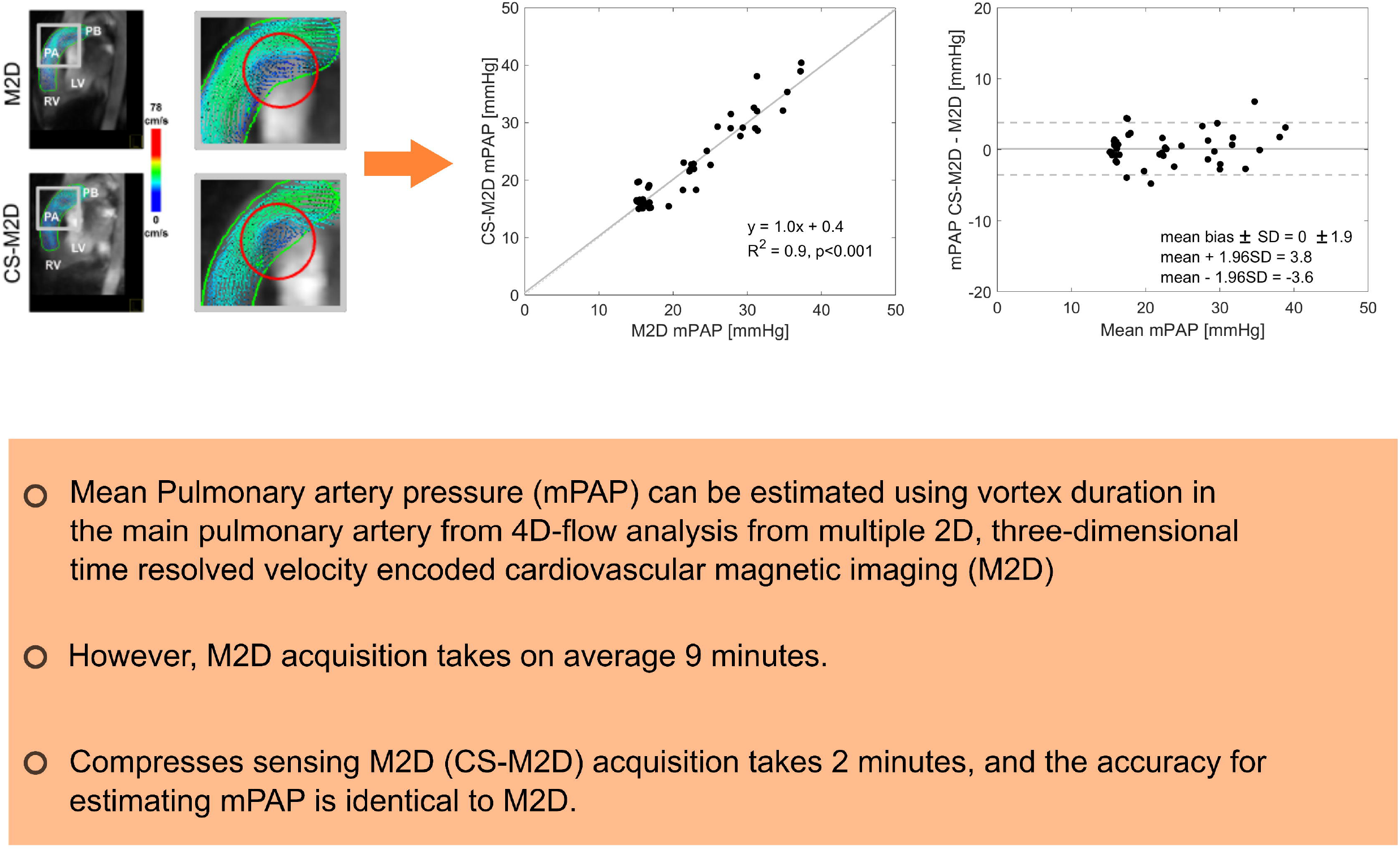
Accelerated flow acquisition for non-invasive estimation of mPAP. Accelerated flow acquisition for 4D flow analysis using compressed sensing technique enables accurate assessment of mean pulmonary artery pressure (mPAP). However, with the important advantage of being faster, which enables the use of CMR 4D flow analysis to determine mPAP and diagnosis of pulmonary hypertension (PH) noninvasively. M2D – Multiplanar 2D, CS-M2D – Compressed Sensing Multiplanar 2D, RV – right ventricle, LV – left ventricle, PA – main pulmonary artery, PB – pulmonary bifurcation. mPAP-mean pulmonary artery pressure.

